# UK health and social care use during and after the pandemic: A qualitative study of the experiences of disabled people from minoritised ethnic groups

**DOI:** 10.1101/2025.02.12.25322134

**Authors:** Carol Rivas, Kusha Anand, Elizabeth Ball, Samina Begum, Neelam Heera, Yesmin Shahid, Zainab Zuzer Lal, Amanda P Moore

## Abstract

The COVID-19 pandemic acted as a “super-catalyst” in accelerating the United Kingdom transition to remote health and social care, while exposing and deepening existing inequities. The 18-month mixed-methods longitudinal CICADA study explored this from the perspective of disabled individuals from minoritised ethnic groups (including undocumented migrants and asylum seekers) during and after the pandemic. Since our aim was to inform improvements, we used an asset and strengths-based approach underpinned by the embodiment model of disability and intersectional theory. Here we report on findings from 271 semi-structured interviews with disabled individuals from minoritised ethnic groups, follow-on workshops with interviewees in April and September 2022, and 4 key informant interviews.

Findings revealed widespread dissatisfaction. Many found the shift to remote care inaccessible and disempowering. Challenges included difficulties in securing appointments, disrupted patient-clinician relationships, little regard for holistic care or comorbidities, and systemic exclusions due to intersecting and discriminating factors of language, accent, disability and complex needs, digital precarity, and undocumented status. Often, service management of expectations would have improved experiences. The increased burden from health and social care support-seeking counter-intuitively worsened once pandemic restrictions eased and led many to give up; others bypassed general practice or raised complaints, or relied on self-management, traditional remedies or informal support networks. Many resorted to costly and unaffordable private care, often specifically from within their communities, and often having never registered with the National Health Service. There is an urgent need for a more inclusive, tailored approach to health and social care that considers intersecting disadvantages of race, disability, citizenship and socioeconomic status, facilitates community connections and empowerment, and provides support for self-care, alternative care and education. By foregrounding community voices, this research offers valuable insights for policymakers and providers aiming to address disparities and improve health and social care outcomes for marginalized populations.

## Introduction

Although the World Health Organisation (WHO) downgraded COVID-19 from a public health emergency of international concern (PHEIC) to an established and ongoing health issue (1) in May 2023, the impacts of the pandemic are still being felt in the UK National Health Service (NHS). One particularly evident impact comes from the way it acted as a “super-catalyst” in accelerating the already planned shift towards remote NHS healthcare, to control COVID-19 infection. This was especially evident to patients in terms of general practice and community care, as the first line of access to the NHS, and the point of referral to secondary care. Prior to the pandemic, around 84% of GP appointments were face-to-face. This fell to 54% in March 2021 in England, with similar trends in the devolved nations (2). The proportion of face-to-face GP appointments then began to rise steadily between 2021 and 2023, before beginning to fall again to 68.4% in January 2024 (3). Digital triage systems were also implemented during the pandemic and remain, e.g., eConsult (2). These were seen as necessary by NHS leaders but an encumbrance by patients, who did not feel they mitigated the tremendous difficulties patients reported in phoning for an appointment. Clinicians also often disliked remote consultations and the attendant processes (4).

The pandemic also highlighted existing care inequities. Public Health England (now fragmented across the new UK Health Security Agency (UKHSA), Office for Health Improvement and Disparities (DHSC), NHS England, and NHS Digital (5) responded by developing a Health Equity Assessment Tool (HEAT) to support organisations to think more systematically about the impacts of their services on these groups.

Among impacts that have not been sustained, there were many reports of bureaucratic “red tape” being lifted and standards relaxed so systems could cope in the difficult circumstances of the pandemic. This included reduced documentation, changes to finance allocations and barriers to partnership working (6,7). These changes made it easier for the NHS and local community groups to work together to share information into vulnerable communities. For example, staff found that joining existing Zoom meetings run by charities or support groups helped to build trust and encourage dialogue and collaboration. Notably, collaboration is entrenched in The Health and Care Act 2022’s approach of Integrated Care Systems (ICSs), place-based partnerships and neighbourhood models.

The 2023 British Medical Association (BMA) COVID review (8) recommends that the NHS should learn from and develop some of the positive impacts of the pandemic. It includes hybrid care, in which face to face care is mixed with remote consultations and asynchronous services (where the patient and clinician are not present at the same time, e.g. sending photos by chat or email). From the NHS perspective, this can help address backlogs in care, and NHS estate limitations, and improve staff well-being and productivity. For patients, it reduces the need for travel to the GP or hospital, which is particularly beneficial for some patients, such those with mobility and transport difficulties or who wish to avoid time off work.

However, the BMA and other recommendations cannot be successfully implemented without considering the perspectives of patients and members of the public in more detail, the issues they have faced since the pandemic began, the legacy of the pandemic, and some of the assets and strengths patients have drawn on as the changes in care were manifest. These considerations were the focus of the CICADA study. We wanted to understand what helped or hindered people in coping with daily life, health and social care during the pandemic, so we could develop guidance and simple interventions to improve these experiences. Our specific focus was on how intersecting factors of disadvantage and discrimination affected pandemic daily life for disabled people from minoritised ethnic groups, aiming to improve their experiences and social, health and wellbeing outcomes as a result.

## Methods

The CICADA study was a mixed-methods longitudinal strengths and asset-based study, framed using embodiment disability models (9,10) and intersectionality theory (11). The term intersectionality was coined by a black woman, Kimberly Crenshaw (11) to explain the way that multiple intersecting axes of power and privilege lead to structural discrimination. Members of the public and patients were involved from the start, with two as co-applicants helping to shape the research questions and design at grant-application writing stage. The study integrated secondary analyses of existing cohort/panel data and a literature review, primary quantitative data collection with 4,326 survey respondents analysed and invited to complete the survey twice more over 18 months, and qualitative data collection and analysis.

Here we report on health and social care experiences from the qualitative data, which focused on people of Arab (Middle Eastern and North African), South Asian, African, Central/East European and White British heritage with and without disability. We considered as a ‘disability’ any incurable condition or impairment that affected activities of daily life, categorised according to six disability impact categories, adapted from UK Government Statistical Service harmonised data recommendations (12).These were stamina (encompassing all conditions affecting lungs, heart and fatigue), manual dexterity, mobility, mental health, cognition (including brain damage, dementia and neurodivergence), food-relevant and sensorial. We later added two more on the advice of our advisory group (‘cancer’ and ‘brain hyperexcitability’ [epilepsy or migraines]). We included self-diagnoses, to be inclusive of conditions that typically take years to be diagnosed. Initially we recruited across six sites of England (Manchester and the Northwest coast, Yorkshire, London, Southeast England, Newcastle and Cumbria, and the Midlands), but later included participants from Wales and Scotland who contacted us (we had no contacts from Northern Ireland or the Republic of Ireland). We aimed for a range of citizenship states but with targeted recruitment of undocumented migrants and those on temporary visas. We used convenience purposive quota sampling, with a goal of 5 participants being interviewed per combination of ethnicity and disability category. With top-up funding we increased this to up to 7 per combination, with a maximum of 294 semi-structured interviews.

Recruitment involved the use of posters, advertisements via our networked organisations and social media. Recruitment and interviews were undertaken by the core team of experienced researchers at University College London (both female, one being an Indian migrant, one White British), our main community partners (Born in Bradford, and Bromley by Bow Community Centre) and eight trained community co-researchers, local to our recruitment sites, who were economic migrants and asylum seekers, some of whom had chronic conditions themselves. All participants provided fully informed written consent. All qualitative data sessions were recorded and transcribed if this was consented to. Community co-researchers underwent training, and their first interviews were checked for quality and rigor, with feedback provided. They were paid pro rata according to the funder guidance for all work undertaken (including training, transcribing and translating where there was sensitive data such as from undocumented migrants who did not want any recordings shared with the central team). Interviewees received £20 per interview. Some co-researchers helped us through the study, with one being a co-author on our final report to the funder; some are co-authors of this paper. We trained our Public Advisory Group (PAG) to became additional co-researchers through all study stages except that they did not undertake interviews. We obtained local and national ethics approvals (UCL IoE REC 1450; Covid-19/IRAS 310741), paying particular attention to anonymisation and confidentiality given that we were foregrounding protected characteristics such as race and disability, and that some participants were undocumented. Raw data, when available, were stored on a secure UCL Data Safe Haven until data cleaning had been completed and were then deleted.

Recruitment used social media, posters, adverts, and snowballing as well as our various networks, partners, community co-researchers and Clinical Research Networks. Through these diverse approaches we tried to include those who lacked resources or technology to respond to online recruitment and people who would not trust the core team because of both historical and current structural discrimination. Participant information materials were piloted within relevant communities.

We held two knowledge exchange workshops with selected interviewees (those for whom we had details, whose interviews were completed by December 2021, and who fitted our core criteria) approximately five and ten months after their interviews (in May and September 2022). We offered face-to-face workshop sessions in each core region or online sessions across regions. To be inclusive, we also offered repeat interviews. These aimed to co-create interpretations of the data using illustrative vignettes recorded by our PAG with verbatim interview extracts. The two sets of workshops also explored subsequent change, assets and strengths, issues and potential solutions; our 10-month workshops used design thinking tools to structure these discussions. PAG members and co-researchers helped facilitate and ensured the workshops were accessible and inclusive. Workshops were followed by key informant interviews (aiming for 15). We also undertook co-design workshops in which patients, community groups and health and social care professionals worked together to develop rapid-impact solutions to issues, and a theatre show, both of which are being reported in separate papers.

The main analysis utilised the Framework approach (13), with deductively determined themes augmented by inductive themes and with data managed in NVivo12 (QSR/Lumivero) and exported into Microsoft Excel^TM^. Full details of the methods used may be found in our published protocol (14). We ensured credibility (with member validation in workshops and an inter-rater reliability exercise between three core team members with discussions and coding of new transcripts until we achieved values of 75%+ for the key themes, as well as our participatory work), transferability (with full description, triangulation with national data and data from the CICADA study’s other strands, and sensitivity analyses), confirmability (using illustrative data extracts) and dependability (via transparent methods and archiving of anonymised data). In reporting our data, we mostly avoid precise numbers and instead indicate approximate frequency using the words ‘a few’ or ‘some’ to represent fewer than 10% of respondents, ‘several’ for 10-24%, ‘many’ for 25-49%, ‘a majority’ for 50-74% and ‘the majority’ or ‘most’ for 75% or more.

The first 210 interviews were completed between 1st July and 20th October 2021. Following top-up funding in spring 2022, a further 64 were undertaken between 1st May and 15th September 2022. Two interviews could not be used because the consent forms were unclear and one because the interview was accidentally deleted before transcription. All remaining 271 first interviews are included in analyses.

### Findings

Interviews lasted 25-90 minutes, workshops took 2 hours. Pre-screening resulted in 40 people being gently told they were ineligible, In total 80 of the 271 analysed interviews were undertaken by partners and lay researchers (almost all face to face). Ten were not held in English and were translated by the community co-researchers who undertook them. We could not quality check the translations as the original data were deleted without being shared with us, at the request of the interviewees, some of whom were undocumented. The central team undertook 30 interviews face to face, four by telephone and two by email; the remainder were via Zoom^TM^. For three interviews, researchers were asked to make written notes instead of recording.

Detailed demographics of the interviews are shown in Table 1. We recruited across most planned combinations, missing only African or Central/East European participants with sensory loss. Our samples were broadly representative of national data (15,16), including the proportions with multimorbidity (17). However, we had fewer participants aged 55 or over. This may partly reflect our heavy reliance on the internet for interviewing due to pandemic restrictions although a third of interviews were face to face. There were slightly more females than males. In our analysis we used a multi-morbidity approach; this uses a holistic perspective when considering the experiences of a person with multiple conditions since one condition does not usually dominate experiences across all contexts (18).

**Table 1:**
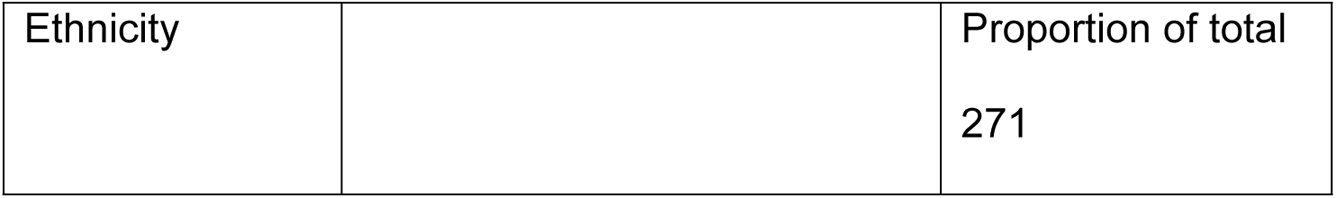
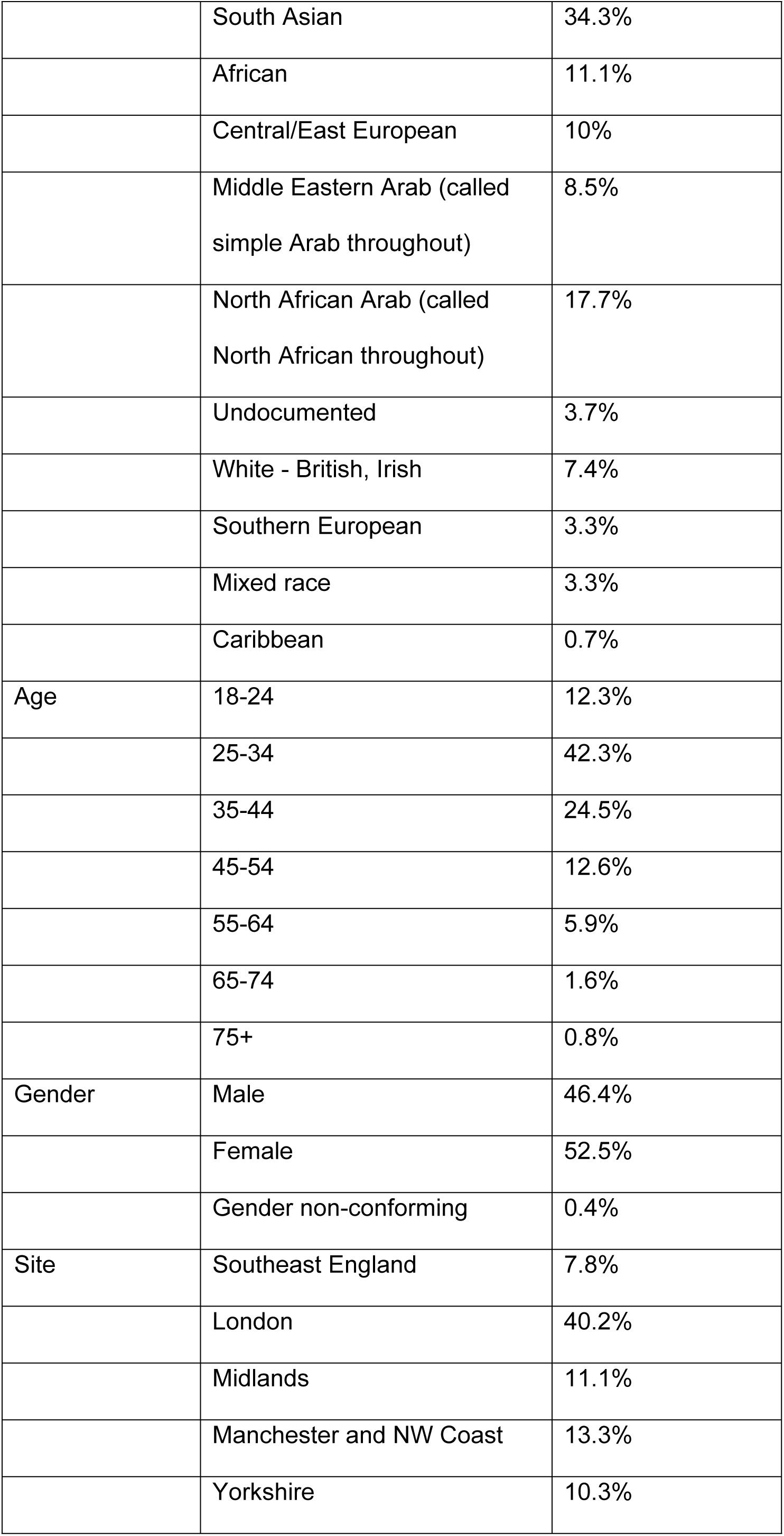
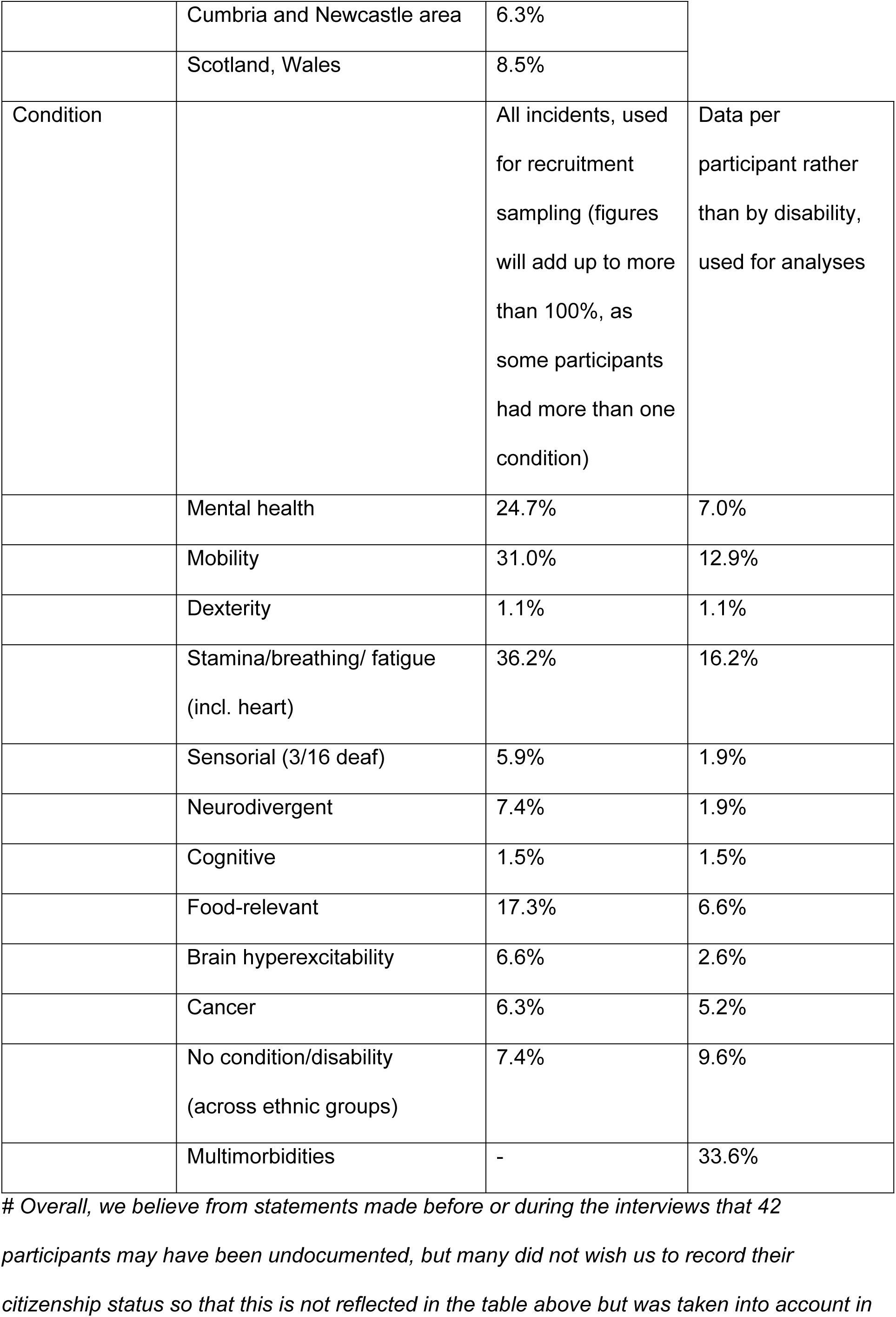
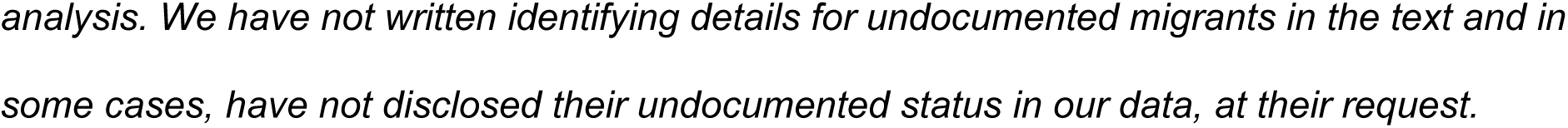
Public participant demographics (n=271)^#^.

Given our intersectional approach, we embraced fuzzy boundaries that challenged our initial categories and so we included data collected on ethnic group identities and sites that fell outside our initial sampling criteria. For example where it transpired during an interview that a person’s parents were from different ethnic groups (a fuzzy boundary) we still included them. However, we conducted a sensitivity analysis, by comparing these data with those that met our initial sampling criteria (our core data) to assess whether our initial sampling decisions were critical.

Altogether 134 interviewees were invited to our workshops; 104 attended in May, 35 in September with refusals in September citing lack of availability due to a return to pre-pandemic routines. We only recruited four key informants, due to the then-prevailing UK political instabilities: a general practitioner (GP), a community leader, a member of parliament (MP) and a charity representative, all from study sites.

### Themed findings

A few participants acknowledged that health and social care professionals were doing their best under the circumstances and 23% praised at least some aspects, either because the NHS had treated their condition as a priority or because of the efforts of individual GPs or local primary care hubs. However overall, our analysis revealed an overwhelming dissatisfaction with health and social care. This is encapsulated in the following themes, described in more detail in the remainder of the paper. They are descriptive to enable integration with the other methods strands of the overall study and policy recommendations.

1. Making the move to remote care
2. Navigating power differences to get GP appointments
3. Disempowered patients and the failing clinical relationship
4. Patient exclusion from appropriate care
5. The use and giving of informal support
6. Alternatives for self-care.

Themes 5 and 6 focus on assets and strengths.

### Making the move to remote care

The move to remote care was mostly criticised by participants, for reasons explored in subsequent themes, but a few specified advantages once they got used to it. As the British Medical Association (BMA) and others have reported (8,19,20), some found it more convenient and a more efficient use of time. We also found benefits not discussed in the literature. This included the reduced need for anxiety-inducing social interaction or travel logistics for some (though not all of those) with neurodivergence, mental health conditions, fatigue or mobility issues or caring responsibilities, or those who were simply afraid of COVID-19 infection. For example, neurodivergent people often get sensory processing overload when in settings where they cannot control one or more from among temperature, light, sounds or smells. In remote care they said they could remain within a familiar environment over which they have full control; they could moderate sound levels or mute the conversation if they need to. Remote care also meant autistics and others with social anxiety or who had withdrawn socially, perhaps because of mental ill health, could avoid the distressing journey to the clinic. As another example, attention deficit hyperactivity disorder (ADHD) can manifest in poor time estimation (time blindness), which can result in chronic lateness or missed or double-booked meetings and appointments. This could be mitigated by clinician calls to the home, as noted by the team in analysis discussions. Nonetheless, remote care can also have disadvantages for these groups, as described in an article by three neurodivergent UK doctors (21).

Participants reported that remote care also allowed parents and caregivers access to medical care for themselves or the person they care for from the comfort of their own homes or offices, and saved the time normally lost by travelling to the clinic and waiting for an appointment. This is especially helpful for those with busy schedules or who work, or who would find it challenging or expensive to bring the person they care for to the clinic or live far away from healthcare facilities. Furthermore, it facilitated discussions of intimate matters for some, which is contrary to findings in other studies (e.g. 22), though for others it complicated these. Others liked the NHS app as a paperless system holding all test results in one place. These benefits were felt across the ethnic groups.

> The last [blood tests] I had the other week was at the London. So he does it all on the system, you haven’t got to walk along there with a bit of paper. So I just turned up, took a seat and waited. (P228, White British, Fatigue/Mobility)

> My unit gave me remote support. My nurse called and sometimes if I needed physical support - like video calls for meals that would be provided […] otherwise, I would have had to have gone in as a day patient. (P9, South Asian, Food-relevant/Cognitive)

Our GP key informant also saw efficiency benefits from a hybrid of face-to-face and telehealth work for patients and in terms of his own time management (“Y*ou can move your calls around between those face-to-face patients*”) but noted he was influenced by the relatively young age profile of his catchment.

> A lot of people prefer to be on the phone because it’s just easier to work around in normal day-to-day life. They could be in Sainsbury’s waiting for you to call them or taking their kid to school or something. They don’t want to be in the surgery. More often are people kind of annoyed that I want them to come in, than are relieved. We just bring them in for a quick examination if we haven’t got what we need to from their history. (GP informant)

Some participants continued to attend GP, secondary and allied healthcare and social care appointments in person. This occurred in our data when critical, live-saving, care was needed, such as for a heart attack, or X-rays and specialist tests or if they had cancer or suspected cancer. Breaking bad news, and some diagnostic needs (such as checking for a malignant melanoma) also necessitated face to face care. In some cases, it also occurred when participants needed routine testing or monitoring or to receive routine intramuscular or intravenous medication. A few had home visits, if they were older or had mobility issues, or cancer. Nonetheless such face-to-face sessions were sometimes reduced compared with pre-pandemic frequencies or alternated with telehealth sessions; this hybrid model has continued to date.

In the heart of the pandemic, in 2021, 44 participants across ethnic groups said they avoided face-to-face care when offered it because they were afraid about catching COVID-19; 16 of these had multimorbidities that particularly increased their risk. These participants managed by:

a. Stopping all healthcare access
b. Only using remote care though needing in person care
c. Modifying their medication intake to reduce visits: *(I tried to take medications, dosages that could last me longer. So, I didn’t have to book appointments to go and get my prescriptions. (P265, South Asian, epilepsy)*
d. Getting the vaccine
e. Using less busy services as routes into needed care:

> My wife did a bit of research for me and so we went to a GP unit. There was hardly anyone there, we went in first thing, they took a look at my ECG and told me to go the Ambulatory Emergency Unit straight away. So I didn’t have to wait in the long A&E with people coughing and spluttering. (P238, White British, Sensorial)

There appeared to be regional and local variability in GP access; though our data could not show this clearly, patients suggested it anecdotally. For example, many participants interviewed in 2022 were frustrated that the GPs of people they knew had fully resumed face-to-face consultations but theirs had not and that ‘the rest of the country’ (i.e., all other services, including secondary care) had returned to face-to-face delivery (‘*Why can’t we see doctors like before, it’s like they don’t care about us.’ P 167, South Asian*).

> My GP practice is working only over the phone […] neighbour […] she has a different GP practice from different area of Bradford, and she said she is able to have an appointment face-to-face. (P175, Central/East European, Stamina/Mobility)

In our April 2022 workshops, participants noted that continued restricted access to services and healthcare was a particular challenge for disabled people. They recognised some benefits of a hybrid online/face-to-face system, but the prevailing processes did not work for most. Some suggested healthcare staff reduced in person contact time with patients because of their own fear of COVID-19 rather than for the benefit of the patient; some GPs were said to only see patients who were vaccinated or could prove they currently tested negative for coronavirus. GP practices operate as independent contractors embedded within the NHS as quasi-employees, and so can make individual choices about care delivery, while other local service delivery may depend on what has been locally commissioned.

### Navigating power differences to get GP appointments

The various challenges involved in making appointments highlighted power differentials, whereby the professionals determined how to handle calls to them, what conditions or situations should be considered more urgent on triage, and when exactly they would call their patient back. This was further complicated by trust and stigma issues, whereby within some community groups, there is an element of secrecy in sharing medical issues as they may be seen as embarrassing. This means necessary information might not always have been presented at the outset to ensure effective triaging.

Twelve percent of interviews completed by December 2021, across all groups, revealed difficulties making GP appointments. While this sounds a low number, frustrations were more common in later interviews and in the workshops with the same interviewees five and ten months later, when participants realised the situation was not temporary. The main challenges are shown in Table 2.

**Table 2.** The challenges of making GP appointments by phone as reported in our data.

| Challenges | Example data extract |
| --- | --- |
| A failure of GP practice staff to answer calls or long call waiting times, which patients interpreted in different and critical ways. This also brought into play technological issues such as dropped calls. | *It's been so bad that I couldn't get an appointment with my GP because the lines open at eight and you call at eight o'clock you're number 29. And by the time they get to you, because you've been on the line for so long the line cuts off automatically. So you dial up again and then you're number 30 in the queue. And then it's just a vicious circle. (P211, South Asian, Food-relevant/Mental Health)* |
| Gatekeeping by receptionists, once they answered the phone. | *And I said it's a Friday, if the doctor doesn't call me in time the chemist will close so I won't get my prescription. But they don't care, it's just the receptionists, wrong people are in the wrong job, they have no empathy, they don't care they're just there to do their job and clock off and finish. You know, they're not even flipping qualified and the way they interrogate you is just like what the hell. I don't want to speak to you lot, I know more about my condition than* |
|  | *you ever will. I just want you to [...] I want an appointment with the doctor. It's a simple request but it's just so difficult now.*<br>*(P203, South Asian, Stamina)* |
| Interpreters not available. | *Because of the language barrier they said they will call me back with an interpreter, and they did after an hour or hour and half, but then there was no available appointment left by the time."* (P95, Central/East European, Stamina/Mental Health) |

Some tried for a month before succeeding. A few said family members or other health services intervened to get them appointments, others gave up (“*the palaver of going through and getting that done is just, I couldn’t do it”,* P16, South Asian, stamina/mental health) and self-medicated, bypassed the GP or otherwise took care into their own hands.

> I wanted to make an appointment at doctor’s….. I was trying it for one, two week. Then because I struggled, I medicate myself and start using medication what I had left. (P72, Central/East European, Mobility)

> One side is saying you have to save NHS money and do not go to A&E, but on other side GP do not want to give you an appointment […]. After they are surprised we are going to emergency department. (P175, Central/East European, Stamina/Mobility)

Consequently, urgent needs were not met appropriately or in timely fashion, with adverse or potentially adverse effects on health. Solutions participants suggested were: callback systems; more phone-in times; more individualised appointment windows; better understanding between GP staff and patients.

GPs set up online booking systems as an alternative to phoning in, which 16 participants complained about. Many others were unaware of this or lacked appropriate technologies as noted in other studies (23). Several reported the online form was tedious and difficult to fill in. It excluded some by language and others by condition, not just in terms of accessibility but also because conditions did not map well onto questions. Some older people felt disempowered in having to seek help from their children. While phone-in booking could result in same-day appointments, online triage led to 2-3 day waits. Participants expressed a need for a revision of triage systems, with alternatives for those who are excluded by them.

> You had to go fill in this online form that took forever. Questions after questions after questions […]. And for every little thing I had to fill out a form. (P206, South Asian, multimorbidity)

> *I had a sudden rash, my face swelled up, I needed to be seen face-to-face, now I called them, and they said I had to fill in a form, I said look can’t you just put me on the call list they’re like no, no, it’s not a simple yes/no, it’s a lot of details you have to give […] she would not budge I had to do it that way.* (P203, South Asian, multimorbidity)

Our GP-linked charity informant was concerned about digital exclusion associated with lack of hardware, access to Wi-Fi, language and literacy and skills barriers, though their description of what GPs told them sits in tension with this and with participant complaints, suggesting that neither patients nor GPs understood each other’s perspectives.

> I think our GPs’ view is that e-consult in trying to solve a problem has created another problem. One of our GPs was telling me this morning, ‘I had to turn e-consult off because by eight o’clock in the morning you’ve got 50 patients on e-consult already and it just keeps coming. You’ve got two doctors on surgery who are seeing physical patients and then you’ve got e-consult coming through just continually and basically anybody at home on a smartphone or a laptop can just go on to the website and go on to e-consult just sitting in your living room. It’s like the sorcerer’s apprentice. It’s created this thing that is just unstoppable. Anybody with an idle moment can just be on their phone or laptop or iPad at home - crazy. It’s the same for the doctors as having [that] patient in the waiting room. They’ve got to deal with it. It’s just created a monster.’ (Charity informant paraphrasing a GP)

Several participants, particularly women looking after children, and those with precarious income on zero-hour contracts, commented on the stresses caused by GP, community service and social care phone appointments scheduled as simply, for example, ‘in the morning’ or ‘by 5pm’. The impact was affected by intersecting factors of disadvantage, such as a lack of English language fluency or childcare support. Our GP informant conceded that ‘*we need to be a bit more flexible and say maybe have reception ask, are there times they [patient] can or can’t make in that morning’*.

> I was walking with my children on the road and then GP called me to discuss something and then my child started to cry. And they said that they would call in the window of like three hours so it’s not like I could stay at home and wait for it. And then GP got annoyed because I couldn’t hear her properly […] I knew it’s urgent thing so it was very stressful and it wasn’t a nice experience and […] my English is not too bad but then I couldn’t understand what she was saying and it was making me feel stupid and not being able to express what help do I need. (P185, Central/East European, Mental health)

### Disempowered patients and the failing clinician-patient relationship

Once participants managed to obtain an appointment, they encountered further issues that were also disempowering. Telehealth meant they were not in control of:

the duration of consultations,
who they were seen by and therefore continuity of care
the information they could impart regarding their health concerns.

This led to:

damaged rapport with staff, and weak confidence in clinical decisions and care.

These issues have been reported in other studies, from the perspectives of GPs (24) as well as patients (19,20). Clinicians were seen by our participants as having lost empathy or interest in the patient.

#### Duration of consultations

A few participants considered telehealth consultations to be too short, and easy for the clinician to terminate, in accordance with 7% of patients in a US retrospective observational study (19). This was particularly problematic since GPs usually operate the rule that only one issue can be discussed per appointment. Two participants said they could extend face-to-face appointments or talk about complex health needs simply by not leaving the room, which was not possible in telehealth. While a survey and a qualitative interview-based study of US healthcare staff attributed consultation brevity to patient reticence or preference (23,24), our participants instead cited discomfort and being made to feel unwelcome and alienated: “*I wasn’t given the time and attention I would have gotten before. With speaking to GPs, they were a lot more abrupt. (P67, South Asian, Mental Health)”*.

#### No continuity of care

We were often told that it was unusual to be dealt with by the same GP on consecutive occasions, a challenge that patients have experienced even before COVID, but more frustrating during the pandemic because patients were not physically meeting healthcare providers and there was no consistent follow-up and coordination. When a locum or unknown doctor phoned, this could lead to issues and misunderstandings, including discrimination and breakdowns in communication. Similar issues were reported across secondary care and social services.

> *When I arrived from Slovakia [after a brief holiday] and informed them I had a stroke [on holiday], doctor replied to me to go back to Slovakia get treatment […] when I have been here for 17 years […] .I also do not know which doctor I am speaking to, I don’t know his name, when I am asking about names, I was told that is not important for you […] That makes me feel so uncomfortable and I am not sure if I do speak with doctor or not*. (P175, Central/East European, Stamina/Mobility)

This lack of continuity, occurring in the context of already disempowering uncertainty about the professional’s level of caring interest, and the time they had available:

- impeded professional monitoring of changes in a person’s condition or situation – something already made harder through remote care;
- could increase patient anxiety as to the quality of their care;
- wasted time and irritated participants, who had to keep re-explaining everything.

Participants and PAG members spoke about having to repeat their histories each time they were seen, with GPs not reading patient notes beforehand, particularly problematic given the lack of continuity of care: ‘*patients have to be accurate historians of their medical journey’* (PAG member).

> I think I’ve only spoken to the same one twice in the past 18 months or so which has been really hard because each time you have to go over your story again and try to remember things that you need to mention […] There’s so much to think about and also not knowing because it’s a telephone consult how long you actually have. (P79, South Asian, multimorbidity)

#### Imparting information about health concerns

Eighteen participants commented that mobility and pain issues in particular could only really be understood in person by GPs, secondary care or allied healthcare staff because of their nature or the way gestures were needed to describe them. Our GP informant agreed.

> I’ve also had one telephone consultation with rheumatology regarding my Raynaud’s in my hands, which is very difficult to describe, because normally you would show them how your hands are, but you can’t do that over the phone. (P79, South Asian, multimorbidity)

> Just seeing someone’s face if they’re describing a mental health problem, seeing someone’s face if they’re describing the impact of chronic pain on their life, what it is they can’t do. You’re really getting a sense of something that you’re missing when it’s just on the phone. (GP informant)

A few participants also considered symptom description hampered by their health illiteracy, lack of English language fluency or education, or the functional limitations of their condition, and a few were asked to read their own blood pressure and considered this would be inaccurate. A blind participant said:

> “Do you have any rashes on your stomach?” How can I say? I can’t see. So, it’s very terrible and horrible.[…] So not possible for me to do over the phone and […] blood pressure, “buy device and do it at home”, but how can I take a machine if I can’t see? (P5, South Asian, Sensorial)

The complaint was often about review or monitoring of established conditions, which contrasts with the literature, where both patients and GPs considered the problem worse for new or developing conditions (19,24).

Video consultations or the facility to send photos to the clinician were generally seen as minimally better than phone conversations. It was argued that these could distort colour or proportions, especially with rashes, while different skin tones exacerbated issues. Moreover, they were not inclusive, marginalising people with technological or financial precarity. There was a lack of privacy (22–24), as well as distraction from family, and technology issues.

> I’d called the GP and they’d had a video call, and they were like, ‘Oh, hold your arm up to the camera,’ sort of thing. [….] They were like, ‘Oh, there doesn’t seem to be anything there.’ I’m like, ‘Okay, well my arm is throbbing and it’s red.’ (P277, Central/East European, Mental health)

> And many people can’t use Zoom. Especially people who are on benefits, they don’t have the kind of income for them to even pay for their Wi-Fi or have good Wi-Fi. (P2, African, Mobility)

> When they did give me a session with a Zoom call, it helped me in the first session, the second one because I had to be in a quiet place but because of the kids in the house, and you are not thinking, looking at the children, they are talking to you, you’re not focusing, it affects the whole of the household, I think. (P210, South Asian, Cognitive/Mental Health)

Some people turned to their local pharmacies to get information and advice because they were easier to access, and the staff spoke community languages. This is something that has been recognised by the UK National Pharmacy Association (25), amongst others.

### Damaged rapport with staff, and weak confidence in clinical decisions and care

Some participants told us about GP encounters that suggested telehealth made clinical decisions inadequate at best and potentially risky. This is supported by UK and Romanian GPs in other studies using a range of methods (19,20,23). When a participant’s condition was downgraded or denied or where reasonable adjustments were not considered when arranging care, patients felt marginalised or gaslighted.

> [The consultant] didn’t know I was a wheelchair user; I had to tell her. (P2, African, Mobility)

> When I got the call, it was very short cannot explain my problem …I have very bad constipation during a pandemic, but I was neglected by my GP and ended up in hospitalisation where I had an enema three-time, a CT scan and was referred to a colorectal specialist for treatment. (P191, South Asian, multimorbidity)

Seven South Asian and Central/East European participants described being given or sometimes they claimed fobbed off by GPs with medications they did not want because of the different power dynamics and feelings of discomfort with telehealth, or lack of trust; similar findings are reported in other studies (20, 24, 26). As with difficulties booking appointments, this led some – especially with mental health problems and anxiety - to self-medicate differently, or research coping strategies or online videos.

> [on the phone] You can talk with him for 2-3 minutes and you have no chance to tell him all your problems and after he will prescribe you stupid tablets even you did not ask about them. (P175, Central/East European, Stamina/Mobility)

Similar issues were described with other NHS services, such as physiotherapy, COPD rehabilitation and mental healthcare; twelve participants across ethnic groups gave examples of how they felt these unsuited to telehealth.

> And [group therapy] was helpful, but it wasn’t as helpful as they used to be in person, because I feel like a lot is lost when it’s, you know, online, like you don’t see the body language and eye contact like you don’t really feel like they’re speaking to people as much. (P43, South Asian, Mental health/Respiratory)

> I am not always able to do these exercises by myself because nobody corrects me. And when I got physical therapy over the phone, it was horrible. […] Where really nothing can be comprehended. […] I think that physiotherapy must go and see and be and touch. (P278, Central/East European, Mental health/Mobility)

Online video consultations were considered better than telehealth for these conditions by the few who had them. But they were described as less good than face-to-face sessions, which were thought to encourage more holistic care and consideration of intersectional factors affecting experiences and mental health; while staff using telehealth could ask relevant questions to elicit this information they apparently did not. This was considered particularly problematic with social care assessments (*you really can’t tell what the person needs over the phone. P2, African, Mobility)*.

### Patient exclusion from appropriate care

The previous theme considered problems within consultations. But what about those participants who were excluded from some care before they got as far as a consultation, because of UK migration policy impacts, language barriers, cultural and structural barriers, disability discrimination, and digital precarity? We now consider each of these in turn.

#### Immigration and the NHS

All UK migrants have some entitlement to NHS care. As a minimum, all overseas nationals in the UK, including undocumented migrants, can access free GP services, and Accident and Emergency (A&E) care. The Immigration Health Surcharge must be paid to get most visas; without visas, foreign nationals must pay a ‘point-of-access’ charge. However, the rules for migrant NHS care are complex to navigate and understand (27), meaning many people are blocked from care even when entitled to it, or are wrongly charged for it (27), or fail to seek it out because they are unaware of their rights. Moreover, since the type of entitlement depends on level of citizenship status, NHS Trusts must check a person’s immigration status which deters migrants even if they are not undocumented. This is driven by Theresa May’s ‘Hostile Environment’ (rebranded the ‘compliant environment’ by the 2019-2024 Сonservative Government), which aims to drive undocumented migrants from the UK by restricting their access to all public services and public funds, and tracking, detaining and deporting them, creating a “*climate of fear*” (28, p1).

We found that as a result some of our migrant participants avoided healthcare altogether, many had never registered with a GP, using A&E as the gateway into NHS care, and many others used private care whether or not they could really afford it. This was true even when, to protect community health during the pandemic, the Government gave a temporary reprieve to migrants, with the 2020/2021 ‘Everyone In’ scheme. It was also true irrespective of rights. While undocumented migrants often had “*no care from anywhere*” (P78, Cognitive), being afraid of being identified, migrants on visas or with British passports who were eligible for free NHS care still chose private care, because they lacked awareness of free care or trust in the NHS or were loyal to private doctors from their own communities or liked the more personalised approach. In several cases, the private doctor was a friend, family member or neighbour who also worked for the NHS, particularly described by North African and South Asian participants: *“I occasionally talk to a friend or a family member. They are also doctors. (P96, South Asian, Respiratory)*.

> Sometimes he is very understanding and sometimes when I’m in a position to, I usually pay for my visits. (P76, North African, Respiratory, Visa)

> I’m used to the family doctor, and I don’t want to maybe betray him, it’s like betrayal you know you have been with him for like 20 years and you have started accessing other services, so that’s why maybe I am not that curious, I don’t want to know. (P250, African, Mental health, Right to Remain)

Some obtained A&E point-of-access free care, then paid the same clinician for private care thereafter. It is unclear whether this was billing for services or a special arrangement, but descriptions suggest the latter. Sometimes NHS staff wrongly interpreted the immigration rules, pushing participants to private care; COVID-19 care was a charge exemption:

> I had some breathing difficulty [because of my lung condition] and my parents took me to the hospital. They refused to take me saying I had COVID, so my parents had to look for a private clinic. The private clinic was helpful. They’re in business, yes, it’s about money. (P25, North African, Respiratory)

Private care from clinically qualified community members (working independently rather than allied to specific community grassroots groups) was viewed positively across groups. As well as offering conventional therapies, these doctors provided clarification and motivation and unlike private clinicians within the broader national system, they met the patients typically every 4-7 days online, and every 2-4 weeks in person, which was liked by participants. Whilst face-to-face care was preferred, private care participants did not complain about remote care, unlike NHS patients, and seemed largely satisfied. However, 34 from minoritised ethnic groups (mostly African, North African, Arab and undocumented) described how private care strained them, since they were in economic precarity, which frequently meant difficult choices about which care to pay for, or premature cessation of essential care. Often family members or friends helped pay.

> There are some things like medications and then there are supplements.[…] CT scans X-rays I have to pay the doctor to get through all those […] there was [mental health] support, but I didn’t go for it because I was discouraged because […] I didn’t have the financial if maybe the therapist or those people need me to pay them I couldn’t. (P264, Arab, Cancer)

A few used doctors in their country of origin, by phone or online, some of whom sent medicines to the UK, which were delayed during parts of the pandemic (“*during the pandemic that became more problematic […] so I resorted to informal acquaintances, and I just asked them”. P19, Arab, Food-relevant; “In 2020, the medicines took time to come to me. I had to wait for a month for medications which affected my health.” Undocumented, food-relevant).* One lady, on a long NHS waiting list, returned to Poland since private care was cheaper there. She could not sustain this, so her problems returned.

> Then at the first lockdown I was in Poland […] I went there and made a private appointment with a lady for physiotherapy, and I had to pay because here in England it took too long and over the phone. […] in England it was only privately, and it was cheaper in Poland. Airline tickets were cheaper during COVID, so it was more profitable for me to come to Poland. So, I flew once a month for two weeks [at a time]. I rinsed out of all my savings. It helped me and I thought I would go back to work but because there is no continuation it gets worse, and everything comes back. (P184, Central/East European, Mobility/Mental health)

One talked about a doctor in her country of origin and a ‘doctor in a garage’ here, an NHS doctor working privately. She describes the double bind of not being allowed to work or have free NHS care but having to work to pay for the private care she is forced to use. Our politician key informant lamented similar cycles of need in other undocumented migrants.

> Then I went to my doctor [the private doctor] in [area of UK] for medicines. They charged me £60 for 10 minutes. It is expensive for me, but I have to ask for his help. He gives me medicines and checks my eyes, but I have to work hard to pay him for every visit. His clinic is in a garage in [town]. It is a small room, and he has a bed and medical instruments to see a patient. (Undocumented)

Our GP informant bewailed: “*They come back with test results that may or may not mean anything and drugs that we may or may not be licensed to give in this country*.” He provided an alternative perspective:

> People are paying for really inappropriate tests that private medical care will give them, and the NHS probably wouldn’t. Not because they don’t need it but because it’s not indicated. I almost have to beg some people, who I know don’t have the money, not to waste it, because it is not a fruitful avenue to pursue in terms of finding a health problem. So, if they really think they have brain cancer, I will send you for a scan, but you don’t have brain cancer. Occasionally you have to do stuff that isn’t indicated on the NHS, as long as it’s cheap enough, and you’re not sending them for MRI scans and expensive things. It is going to create a problem for people who do need it. Just to convince them they don’t have a health problem that they claimed they did to start with. That happens a lot, and people in Eastern Europe, and particularly Nigerian, but also Indian and Pakistani families will, ‘I’m just flying back to get some medical care.’ You’re literally in a country with free medical care, why would you do that? Of course, it’s mixed in with seeing their own family and doing other things, but they’re still spending a lot of money on healthcare when they needn’t. (GP informant).

#### Language barriers

Many CICADA participants described language issues in primary care (e.g., accessing care, describing symptoms) (see also Finlay, Hopkins and Benwell (29)), that led them to go straight to Accident and Emergency (A&E) where they could encounter the same issues. Flagging of communication needs only happened late in some participants’ patient journeys:

> My first session with the [mental health] therapist I have online. She was very nice and asked me because I explained that some of my troubles come from the fact that I can’t communicate everything what I want to say in English because this is not my first language. And this is part of why I’m anxious about communicating institutions because it’s hard for me to communicate. So, she asked me if I would prefer to have therapist that speaks Polish, so this was the moment when it’s like, all right, there are some open doors. […] But again, I had to go through all this part when there was no one speaking Polish, or no one was wanting to speak Polish. (P185, Central/East European, Mental health)

Our seven Roma participants all described the complication of interpreters speaking a different language to them. The first extract below shows GPs apparently falsely assumed all Roma spoke one language, which led to errors compromising care. The second extract shows similar issues with language dialects, in this case Arabic.

> In my experience the interpreter explained my health problem to the doctor the complete opposite […] I received an invitation from GP for a head examination, but I never had a problem with my head. I went to the doctor for a back examination […] they sometimes give us interpreter who speaks Slovak. Slovak is similar to Czech but not the same as Czech language. Therefore, interpreter in Czech should be available for the Czechs nationals. However, the best solution would be an interpreter from the Czech Republic who speaks the Czech language but also the Romani language, so that communication with the patient from the Romani community/nation is improved and inaccurate data towards the doctor do not occur, as this is a complication and could escalate medical appointments. (P72, Central/East European)

> They need to […] use translation from the same community.[…] They used to get me Arabic speaking translators who speak differently. They could not understand my language and I did not understand their Arabic accent. (P87, North African)

This led to mistrust, so some participants wished they could use or resorted to using family interpreters, against NHS guidance (30): *“Fortunately, I have a son […] he communicated [for me]. Doctors offer translators but those cannot be trusted. They distort reality. (P72, Central/East European, Mobility).* Our GP informant, while acknowledging the issues with family interpreters, suggested recommendations to not use these should be balanced with patient needs:

> On one side, you think it’s inappropriate [for children to interpret for their families] and you don’t know really what you’re getting out of it, and the parent doesn’t want a formal interpreter because some of these communities are not huge, and it might be someone that they know or a friend of a friend. On the flipside, some of these children are astonishingly capable young children. (GP informant)

The problems of hospitalisation with no English and no interpreters were exacerbated during the pandemic when interpreters were over-stretched and multiple family members might be ill at once, or when family was forbidden from accompanying patients. At best this meant patients and their families lacked information (contrary to the NHS Act) (31). At worst it could lead to deteriorating illness and even death.

> My mother-in-law and father-in-law [suffered badly from COVID-19 and] were hospitalized [.…] My mother-in-law has been in hospital for nearly 2 months [.…] If I could not speak this little English as I do, we would know [.…] nothing about them. I tried to translate between them. We were only able to get an interpreter at one time at this whole time, and it was still difficult to get one. So, I tried to translate, however I do not speak English very well [.…].The pandemic made me realise how much [interpreters] needed. As we needed them for more than one member of the family at the same time. (P95, Central/East European, multimorbidity)

> She called an ambulance [because she had COVID] [.…] UTC (Urgent Treatment Centre) was really, really busy she was left to sit on the wheelchair outside, but she was on her own not knowing what was happening, very unwell, been waiting for a long time [.…] because of her language barrier she couldn’t ask for help, or she couldn’t go to reception, she ended up crying and then a passer-by gave her his phone and she called her husband and asked him to take her back. So, she went back home. And then she only just deteriorated, worse, worse and worse and then [.…] she was taken to hospital, and she passed away in March after a long battle with COVID. (P235, South Asian, Fatigue/Mobility, about a neighbour)

Some patients became afraid, not knowing what was happening due to the lack of interpreters (explained by P167 below), a situation worsened by lack of trust in the NHS and circulating myths. Three Pakistani participants specified the same conspiracy theory about hospitalisation, evidently linked to the greater pandemic mortality rate in minoritised ethnic groups.

> I was scared because I couldn’t speak or understand English. They told me I would see a Pakistani doctor. Once he came and explained I felt more at ease (sah pirya). (P167, South Asian, Mobility)

> A lot of people were saying don’t go into hospital, especially the old people [because] they give you a lethal injection and you die. It may not be COVID, but they put that on the death certificate. So, especially in the Pakistani community people were afraid to go to hospital if they were ill. (P176, South Asian, multimorbidity)

Participants also complained of false assumptions when they spoke English:

1. Being ‘othered’ as migrant, with racist discrimination and ghosting when they spoke imperfect or accented English;
2. Having needed language support removed when they were simply conversationally articulate.

This could aggravate healthcare problems and disempower patients.

> When I went to GP practice, I have been told I never had a stroke showed them all my paperwork translated in English and diagnosis are multiple sclerosis and stroke and they said that cannot be true. I had a stroke in Slovakia, and I had to translate my hospital report in English, and they denied, and I said only you are a doctor’s and in Slovakia we have no doctors? I was sent to CT scan, and they did apologise after results from CT scan and said there was a stroke. (P175, Central/East European, Stamina/Mobility)

> I ask my doctor for an interpreter, and I am not always able to get one, because the lady at the reception is able to say that my English is good. Well, maybe blood sampling is good, but when it needs an explanation of some result, from a blood test or something, some medical terms can stress me out so much that you tell me that I know, and I don’t know. (P278, Central/East European, Mobility/Mental health)

Our community worker informant said people chose GPs with language-in-common including idiomatic use of English, who they trusted better, even paying to do so, which adds nuance to our finding that participants eligible for NHS care turned to private care:

> We have in West London a GP from one of the islands. People will save their pension money - particularly for the older people - to go to see him rather than go to their doctor because they do not think [the GP will understand what they say], a lot of GPs. With trauma or distress, very often they can become practically incoherent or the language you use they do not understand. They cannot express themselves, so they use a lot of colloquialisms and a lot of home language. The doctors, they only have a few minutes, so people then don’t go to their doctor. They don’t trust them and they will go to see this doctor that people know. He must be so popular! He sees people privately and then people get a private prescription, but they trust him. Even if what he prescribed was no different to what their GP might prescribe, the point is they go to his catchment area, they’re prepared to pay, they want to be listened to, they want somebody to respect that what they come with is important. The fact they can trust that individual, talking (Community worker informant)

#### Cultural barriers

Very few African participants talked about mental health. Those with Central and East European heritage spoke of not help-seeking because of pride, not wanting to seem a burden on the state (encompassing any condition), or the taboo attached to mental health.

> I’m very self-sufficient so I decline all help. I have difficulties with accepting help. It creates shame and feelings of guilt and inadequacy, so I haven’t ever spoken to anyone about my issues before. (P170, Central/East European, Mental Health/Stamina)

> I don’t think I’m seen as mentally ill enough to get the help, or I don’t portray myself as mentally ill enough, because I don’t wear it on my sleeve, but I think because I don’t - well, I think for an Eastern European culture it’s not very normal to speak about mental health, or to discuss it with people. So I think I’m still in that mindset of if you don’t speak about it, it doesn’t exist, sort of thing. So I think I struggle finding help for it, but I think that’s more of my own issue than an outside factor. (P277, Central/East European, Mental health)

Some participants specified healthcare professionals did not respect these and other cultural norms or appreciate the impacts of culture on help-seeking or misunderstandings about care or patient behaviours.

> Things like the treatment I got was very British, western, and I eat with my fingers in my culture, and I wasn’t allowed to do that because they said that’s an anorexic thing. (P9, South Asian, Cognitive/Food-relevant)

Several said doctors should embrace traditional approaches and prayer in tandem with the medicalised. The triumvirate of God, individual effort, and professional support has been noted in other studies of minoritised ethnic groups (32).

> For my husband for instance […] I would get him the spiritual help, but I would do it as a Muslim. […] But also, medically I would want that intervention as well […]so it’s balanced. And then working with both would allow his healing quicker. Whereas I think previously it was just like medical, medical, well we need to give you these tablets and then people weren’t understanding, no, no, no. (P243, South Asian)

Many participants – particularly those with mental ill health – believed waiting lists were purposely extended for minoritised ethnicities because they were vulnerable to being fobbed off with incorrect information and were considered a nuisance and less important than the white British. Others highlighted as an alternative explanation the structural barriers to access such as lack of knowledge of the system and language problems.

> I’ve been struggling to get any help; I’ve been under the mental health team for three or four years and I’ve not had any help or treatment from them for the last three years. I’ve been on waiting lists. I’ve been waiting. I’ve been chasing. I’ve been complaining. Again, it’s the thing about ethnic minorities. They don’t take us seriously in anything, in the services in this country, we’re just seen as a pest. Or we’re just seen as like a bother, or we’re just seen as like- and if you complain or if you raise any issues, they will just use it against you, or they’ll take advantage and lengthen things out on purpose. (P7, South Asian, Multimorbidity)

> Because I don’t know how the system works, sometimes I can feel that maybe they can just – how to say this – they don’t treat me like an English person let’s say, because I don’t know how to fight for my rights, I don’t know where to go, I don’t know what I can and what I can’t, so I don’t know the system properly, so they can say to me, “Yes we do it like this,” but actually if there would be an English person they know that no it’s not right, you can still do something, that they can do it. (P196, Central/East European, Mental health)

#### Disability barriers

The UK Government Equality Act 2010 places a legal duty on all service providers, and an additional duty on public sector bodies, to make “reasonable adjustments” that avoid putting disabled people at a substantial disadvantage compared to those not disabled. Ironically, we found participants were often unable, because of their disability or condition, to do the therapy or use the equipment provided to help them manage their disability or condition.

> I’ve got deformity in the hand, so I have gripping issues, so I wasn’t able to do that therapy myself. (P234, South Asian, Stamina/Mobility)

> I’ve got a wheelchair. I’ve got nobody to push me. Now it’s a self-propelling wheelchair, but I can’t propel it, because fibromyalgia in both arms. (P64, South Asian, Multimorbidity)

#### Digital exclusion

NHS Guidance on remote communication is weaker than for other aspects of care and does not consider the intersection of language issues with sensorial impairments, age, digital poverty or inadequate digital skills (see also Finlay, Hopkins and Benwell (29)). Our data showed this could exacerbate issues. By September 2022 it was clear from our workshops that the new hybrid, partially digital world increased the care burden of members of minority ethnic communities who were tech-literate with English language skills, supporting elders in the family and wider community - ‘*now we do everything. We are their advocates, their translators’*.

> I don’t know how to use smart phones or email, [ social services] could not help me, no carer, nobody to help with booking appointments, nobody will help me to send emails. They were very useless I want to reach to human rights organisations. I feel that there were violations of my rights. Because I am an old lady from minority group and don’t speak English, I am deaf, so they give up and get frustrated. (P87, North African, Mobility/Sensorial)

### The use and giving of informal support

To manage the different issues, practical support from family and friends was described as of critical importance to many through the pandemic and in its aftermath. It included delivering care normally provided by health professionals, such as weekly physical therapy, and practical assistance for shielding participants and others unable to access medication without help. Recent migrants were often dependent on family and friends for financial assistance to fund medications and care. In multigenerational households, older participants were practically supported by younger family members to navigate online doctor’s appointments and technology and communicate with healthcare teams. Family and friend emotional support was also important across ethnicities and health conditions and potentially prevented some psychological issues, as well as helping those who already had mental ill health.

Community emotional and practical support, via neighbours, charity groups, faith institutions or other informal networks, supplemented family and friend support or compensated for its lack, though some participants lacked community support. Neighbourhood ethnic diversity appeared to influence the degree of connectivity, with participants of minoritised ethnicity living in mainly white neighbourhoods feeling less supported. Charities and informal networks, for example on Facebook, facilitated ties and support between people with similar conditions, and when access to healthcare was restricted, provided lived experience-related advice and other useful condition-related COVID-19 information, as a substitute. Apps like TikTok, YouTube and online courses run through charities and other organisations facilitated self-care. Charities also provided mental health support and therapy. Some with disabilities requiring close therapeutic support relied on a therapist, clinician, formal carer or social worker, although generally ties appeared weaker than for informal networks.

> We got this Facebook Page […] so, if you post a question, they will try to help you or answer your question, maybe someone has a similar experience and then they can guide you. (P12, South Asian, Sensorial)

> We have this organisation where we talk with people, they have some people who are disabled like me, like maybe hands are not working, so we shall come together and maybe try to talk, and exchange ideas on how to solve some problems, and talk of many stories. With that I just feel relieved […] the situation I’m suffering from fades away. (P33, North African, Mobility)

Empowered participants coped better than others; they were able, for example, to support others in the community, advocating for health and social care rights for their self and the community. A few mobilised local support networks and fought for service access. They explained the value of good English, education, and knowing your rights:

> They wouldn’t dare speak to a Caucasian person […] in that manner and then when I’ve spoken to them [the Council] on their behalf, I see the complete tone change and when I say this is unacceptable, can I raise it further, can I speak to the manager and then I get that and then the people who I help they’re like oh thank you so much I know if you hadn’t come and if you hadn’t spoken to them, I would have not got nowhere. (P203, S Asian, Cognitive)

### Alternatives for self-care

Alternative self-care strategies associated with enhanced mental and physical wellbeing were undertaken by 43 people across the core ethnic groups, including most commonly a third of African participants and a fifth to sixth of South Asian, Arab and also White British participants. Strategies included hobbies, mindfulness, yoga, meditation and positive affirmation, and personal spirituality. These might also be offered by formal support services.

> I would say self-help spiritual healing [helped me]. So, I believe in spiritual things, like karma and affirmations, and I’m doing a lot of work on myself, like rewiring, reprogramming. So, building things up to not see things from a one-pointed lens, not take things so black and white. So, I’ve been using the time to kind of work on the foundations with myself. (P9, S Asian, Cognitive/Food-relevant)

Those with financial stability and adequate accommodation used lockdown as an opportunity to focus on their self-care, following traditional recommendations for natural tonics and herbs to boost immunity, taking supplements, eating well and exercising:

> The pandemic meant that I didn’t really have to go anywhere or focus too much on other things because I could do it from home. I actually focused a lot more on my diet and my exercise. Because the only thing you could really do in the pandemic was go out for a jog or whatever. (P26, S Asian, Fatigue)

Traditional remedies appeared comforting, and emotionally supportive as well as being ascribed physical benefits – “*the ginger garlic drink […] is good for my body and immunity […I think this makes me more confident and settled” (P100, African, Cancer).* Walking was the most popular exercise across ethnicities, even for people who struggled to do so without help. People with conditions managed by healthy lifestyles e.g., diabetes, in difficult living conditions walked 10,000 steps in the flat or compound. Walking helped families to bond in lockdown and meet others outside, even though ‘*we would walk 2m apart wearing masks’*. One person said: *“sometimes [NHS help] is not tailored to our condition and experiences in life, so I also started speaking to the gym instructor on how to stay healthy during lockdowns. (P200, S Asian, Stamina)”,*

However, post-pandemic, participants, talking in our April 2022 workshops, found it hard to continue these activities. Online courses were no longer free, they had competing pressures such as the need to earn money, or they needed help with activities and their families had gone back to work. Their stories highlight the potential value of social prescribing.

> I am suffering from the chronic depression from last 13 years […] So, what I started doing is meditation and listening music more and using video calls, that sort of things […] .And doing exercise at home. So, these kind of things helped me a lot. And actually, this COVID thing has helped me a lot. Like it was a blessing in disguise that year, I have lost a lot of weight by exercising at home. I was always shy to go to gym because I was so overweight. Now I am not following the same pattern or routine because the life is coming back to normal a bit. (Leicester)

## Discussion

Our participants struggled with health and social care through the pandemic and into the end of 2022, when the CICADA study ended. Our findings corroborate studies for people with intellectual disability (33), dementia (34–36), and of South Asian heritage (26, 37) but add nuances. There was a lack of holistic care and a disregard for comorbidities and intersecting factors of disadvantage such as social and housing care needs. Difficulties making GP appointments by phone or via e-consult-style triage were exacerbated in those digitally impoverished, with complex health needs or not fluent in English. An informant comment that GPs felt overwhelmed because patients ‘can just go on the website’, encouraging more trivial help-seeking, sits in tension with participant complaints of lack of online accessibility.

Our data highlighted various power differentials, for example with GP triaging decisions dictating the timing of callbacks. Non-specific remote appointment times were particularly detrimental to those lacking income or support networks. Impaired patient-clinician relationships led to perceptions of discrimination, and migrants’ lack of knowledge of UK processes was seen to make them more vulnerable to being ‘fobbed off’ by GPs with tablets without referrals. This illustrated the different power dynamics in remote care, and patient feelings of discomfort with telehealth; similar findings occur in other studies (20, 24, 26). As with difficulties booking appointments, this led some – especially with mental health problems and anxiety - to self-medicate differently, or research coping strategies or online videos. Issues with diagnosis and monitoring, as noted in the literature (24, 26), were confirmed, particularly for some conditions hard to describe verbally or where the patient could not see. Face-to-face care continued for those most needing it. However, several participants – especially with multimorbidities - refused it through fear of COVID-19 (as found in a dementia study (36), stopping all healthcare, only using remote care, reducing visits by self-medicating with reduced doses, getting vaccine protection, or choosing less busy services.

Our findings sit in tension with a systematic review of the literature on remote consultations which focused on the general population (23). We do not support its findings – at least in our index group – of better monitoring of cases, increased insights into the home environment, empowerment to discuss more personal issues (our findings were equivocal). We did however find some similar benefits - convenience, reduced risk of COVID-19 infection - and the same issues of compromised clinical decision-making, privacy concerns, technology barriers, loss of rapport, too-short consultation times, language barriers and difficulty booking consultations.

We were able to undertake some longitudinal analysis. This showed the situation often worsened for our participants as the pandemic wore on and the country opened up. The new norm of hybrid care and online triage increasingly disadvantaged those with mental health issues, sensory disabilities and complex chronic conditions in particular. For example, telephone consultations were no longer possible for a deaf participant as her partner had returned to work, the insistence on same-day in-person appointments meant taxis and carers (such as family members now back at work) could not be planned and organised while those who needed to plan ahead for appointments to ensure they could cope psychologically (such as autistics), and physically (such as people with disabilities that reduced their energy levels) were not catered for. Two years of largely remote care meant lost rapport with primary care teams. People missed one-to-one interaction but also felt hostility from frontline reception staff and felt healthcare staff were ‘desensitised’. There was frustration that hybrid processes were not ‘joined up’, leaving people without medication, and with lost test results. Collectively these factors meant that in late 2022, people were still continuing with strategies they had developed during the pandemic, such as going straight to A&E, seeking private care even when they could not afford it, self-managing care and treatment (often sub-optimally) and on occasion *‘gaming the system’* such as exaggerating symptoms on online triage forms to get an appointment. There was less frustration about secondary care, beyond extended waiting times though feelings of being held in limbo and neglected had intensified in those affected by the care backlog (38). Access to social care was less mentioned, with less reliance on this than during the pandemic, but challenges remained, created by online access and budget cuts.

A new and important finding is that many participants had never registered with a GP, irrespective of residency status. They trusted private care more though they could not often afford it, as it better matched their expectations from their countries of origin. Poppleton (39) has described this for Central/East Europeans; we found they commonly spoke of better systems outside the UK, but this phenomenon cut across the ethnic groups. The financial burden was particularly described by Arab and African participants. Sometimes, the private doctor was a friend, family member or neighbour who worked for the NHS. A few used doctors in their country of origin, by phone or online, some of whom sent medicines to the UK. The issues inherent in these practices remain to be explored though it is important for health and social care services to work alongside existing habits, processes and structures rather than criticising and discouraging them.

Our data also suggest it might also be helpful to further investigate the ongoing hybrid model of care and how it is affecting different groups in various ways. A scoping review of the literature has identified inequities in remote care by age, race, SES, health conditions, eHealth literacy, and geographic location (40). The authors reported that older adults may be unable to use technologies for various reasons even when they are digitally literate, for example because of failing eyesight (40). Nonetheless some inequity gaps are closing (41). Smartphone ownership among immigrants is close to 90% in countries like the UK and US (41,42). However, if phone applications are not designed in the native language of immigrants or with their needs in mind, these apps are less likely to be effective or utilized (42). Better command of the English language has been associated with higher odds of having used digital services among naturalized citizens and noncitizens in the US (43), which is consistent with research on the use of healthcare in general. Obstacles to accessing phone and video appointments include language barriers, either because English is not the first language or because the person may not be able to verbalize their complaints well in remote care, access to phone/video technology or WIFI broadband, and lack of privacy due to domestic overcrowding. Other important obstacles are lack of education and engagement (44). These points emphasise the complexity of people’s experiences and underline the need for more tailored approaches in health and social care for disabled ethnic minorities. It is also important to note that many clinicians found remote care problematic; not only have many reported issues with diagnosis in the literature, they also felt it added what one of us, a practising clinician, has termed moral injury.

### Patient burden and help-seeking

May et al (45) described the increasing reliance of health services on individuals’ self-management of their conditions, making patients increasingly accountable for this work. Overall, we found the pandemic increased this treatment burden leading to a feeling of abandonment by healthcare, increased distrust in formal care, and increased dependence on informal relational networks. This was augmented by fear of COVID-19 and unclear information on what people should do. Similar reports of increased self-management work and linked issues were identified in a study of white British participants, many multimorbid (46).

Shippee et al (47) described a feedback loop whereby increased patient workload from an increase in the burden of symptoms, treatment and everyday lifework caused by the pandemic reduces a patients’ capacity to access healthcare or carry out self-care. This was supported by our data; many participants described giving up on the NHS; others said exhortations by NHS staff to self-care did not consider the constraints of their disabilities that hindered this. They were also worried about COVID-19 infection in healthcare settings. In our parallel survey data, reported elsewhere (48), we found having chronic health conditions could reduce help-seeking from the NHS and increase help-seeking from the community, with social and mental wellbeing benefits by late 2022. One explanation may be that people with established disability who are not in a health crisis will have learned how to self-manage their situation so that they need help for structural rather than specific health barriers. For example, many reported language barriers, interpreters with the wrong language or dialect, being ‘othered’ as migrant, with racist discrimination and ghosting when they spoke imperfect or accented English, and having necessary language support removed because they were conversationally articulate. This would also help explain reduced social wellbeing associated with NHS support, as found in our survey data, which therefore seems to have multifaceted causes.

Our data suggest that local and national policies should focus on facilitating community connections and empowerment, and opportunities for self-care, alternative care and education as these were associated with reported mental and physical wellbeing. Some assets, such as access to technology, and to courses (for hobbies, alternative care or skill development), cut across potential policy strategies and should be prioritised. Others, such as the use of spirituality and traditional remedies, should be better accommodated by formal services, working in tandem with these, for their psychosocial benefits. Walking and dietary considerations were both mentioned by many across groups, indicating the importance of social prescribing. Stories of empowered participants, advocating for others, raising complaints, or otherwise dealing with health and social care issues, highlight pervasive challenges of White British centrism, racism and disability discrimination and the importance of capacity building within communities. It is important when considering these policy suggestions, for structural inequities to be the focus of the intervention, rather than pushing responsibility and accountability back to minoritised communities or referring to their cultural values and practices as supposed challenges instead of focusing on the need for structural change (49). This can be supported by the co-design of services including underserved patients, which this study has shown can be done. Indeed, we are developing some of the ideas generated by our study for local and regional implementation.

### Intersectionality

Our findings on the intersection of citizenship status with disability in worsening access to healthcare accords with other work on asylum seekers specifically (29, 50), but we have a broader range of citizenship states in our data. We found citizenship status also intersects with socioeconomic status in terms of income, employment prospects and accessing private healthcare during the pandemic. We found intersections between ethnicity and mental health in perceptions of being fobbed off by health providers, though as with all intersecting relationships we report, the very nature of intersectionality means many other intersecting factors were also important in terms of structural discrimination, such as language fluency and degree of assimilation in the UK. We found language, culture and even disability were often not taken account of in medical care, and so this could exclude, or cross religious and cultural lines, or result in inappropriate and potentially harmful intervention, or destroy clinician-patient relationships and lead to perceptions of discrimination. Our data confirm other work (32) showing that religion, culture, and the biomedical should be harnessed in tandem. There was a lack of holistic care that considered intersecting factors despite common co-existence of disabilities and social and housing care needs. Non-specific appointment times for remote consultations were particularly detrimental to those with combinations from among low income, disability, lack of support networks, childcare needs and English language fluency. In many cases, service management of expectations would have improved experiences. NHS Guidance on remote communication is weaker than for other aspects of care and does not consider the intersection of language issues with sensory loss (e.g., deafness, sight problems), age or inadequate digital skills. Our data showed this could exacerbate issues, and e-triaging was especially problematic for those with digital poverty, multimorbidities, lack of English language fluency, as well as some specific disabilities such as blindness, musculoskeletal conditions, neurodivergence. While the intersection of different minoritising factors tended to worsen experiences, participants with more disabling conditions mostly focused on disability discrimination only in their narratives.

### Strengths and limitations and methodological issues

Ours is, so far as we are aware, the most in-depth study of the health and social care lived experiences of people from minoritised ethnic groups with disabilities. We undertook 271 interviews and invited over half of interviewees to two series of workshops to consider change at approximately six-monthly intervals. This paper provides perspectives on both pandemic and post-pandemic experiences. Community co-researchers and partners made an important contribution in increasing our reach and the inclusion and engagement of undocumented migrants and others with precarious status, whose voices are normally not heard. We recruited across a wide range of disabilities and ethnic groups, larger than is usual within similar studies, without compromising on depth, and used an intersectional approach from the start.

Nonetheless there are some limitations. Our interviews, mainly conducted online, may have excluded some older and more disabled participants, and undocumented migrants, notwithstanding that co-researchers and partners undertook in-person work in their local communities. However, some participants were from the oldest age groups, significant numbers were undocumented, and we had a nationally representative proportion of people with multimorbidities. We had considerable drop-out between the first and second workshops. We had formal translators/interpreters in only two of our six sites although our co-researchers ensured that consent was fully informed across all sites. Co-researchers translated interviews undertaken in other languages than English and because many were from undocumented migrants or those mistrusting of organisations, could not share the audio recordings of these, so we could not do quality checks on translations. We focused on England though we have limited data from Wales and Scotland showing similar accounts and our triangulating survey (not reported here) was UK-wide (48). Our qualitative data have stronger representation by South Asians than others though we corrected for this in analyses. Recall of events prior to interviews (which, due to top-up funding continued into summer 2022), may have been inaccurate. However, accounts of earlier and later participants corresponded and aligned with the literature. Further detail on coping strategies and support are discussed in other CICADA papers rather than here, where the focus has been on challenges and empowerment.

## Data Availability

Anonymised data are available from the author for education and research purposes on request.

## Acknowledgments

We are grateful to all the participants who took part in this study, and our community co-researchers and partners who enabled or facilitated workshop events and undertook some interviews.

## Data sharing

The full data cannot be shared publicly because they include responses from undocumented migrants who have refused this permission. The anonymised qualitative data from interviews and workshops will be deposited for archiving and reuse under a Restrictive Licence according to UCL protocols existing at the time. The Restriction is in place to protect the identities of participants who have multiple protected characteristics described in the data and to avoid anonymisation being breached. These data will be available on request for up to 25 years after end of the study (i.e. up to 2047). Archived data will be checked for anonymisation before sharing; raw data will never be shared but will remain in the UCL safe haven. Data that are considered by the custodian to be sensitive and not in the public interest will not be shared despite anonymisation. Other anonymised data will be shared according to a Restrictive Licence and extant UCL protocols. The custodian of the data to whom requests may be made is Professor Carol Rivas; requests may also be made via.

## Ethics

We obtained local and national ethics approvals (UCL IoE REC 1450; Covid-19/IRAS 310741). All participants provided fully informed written consent.

